# Social Determinants of Health Influencing Colorectal Cancer Screening Behaviors Among Community Residents in China: A Scoping Review Protocol

**DOI:** 10.1101/2025.05.16.25327109

**Authors:** Fen Xu, Yang Wang, Lulu Wang, Hua Jin, Dehua Yu

**Affiliations:** Department of General Practice, Yangpu Hospital, School of Medicine, Tongji University, Shanghai, China; Shanghai General Practice and Community Health Development Research Center, Shanghai, China; General Practice Research Center, School of Medicine, Tongji University, Shanghai, China

## Abstract

**Background:** Colorectal cancer (CRC) has emerged as a significant public health challenge in China, with incidence and mortality rates continuing to rise. Although CRC screening has been well established as an effective approach to reducing disease burden, participation among community residents remains suboptimal. This is especially true for high-risk individuals, among whom adherence to follow-up colonoscopy is notably low. International evidence indicates that social determinants of health (SDoH)—including education level, income status, and accessibility of healthcare services—play a substantial role in influencing screening behaviors. However, in China, a standardized framework for the classification and assessment of SDoH has yet to be established. Existing evidence is fragmented, making it difficult to comprehensively understand how these social factors affect screening participation within the local sociocultural and health system context.This scoping review aims to identify and mapSDoH that influence CRC screening behaviors among community-dwelling adults in mainland China. It will also summarize how these determinants are defined and measured in the literature, Provide a foundational overview to inform future research,and equity-oriented screening interventions within the Chinese context.

**Methods:** This scoping review will be conducted in accordance with the JBI methodology for scoping reviews. The literature search will cover both English and Chinese databases, including PubMed, Embase, Web of Science, Scopus, Google Scholar, CNKI, and Wanfang Database. The inclusion period spans from January 1, 2012, to April 23, 2025. Eligible sources will include quantitative studies, qualitative studies, and mixed-methods research, encompassing peer-reviewed journal articles, conference proceedings, and grey literature.A three-stage search strategy will be employed to ensure comprehensive coverage. Two reviewers will independently extract data from eligible studies. The findings will be initially categorized based on the five key domains of SDoH defined by the Healthy People 2030 framework, with additional attention given to China-specific contextual factors. Data to be extracted will include the type of SDoH, frequency of appearance, relevant stage of screening (initial or follow-up), direction of influence, study design, conceptual definitions, and measurement tools. Results will be presented through narrative synthesis and structured tables to support clarity and applicability in primary care and public health practice.

**Discussion:** To our knowledge, this will be the first scoping study in China to systematically apply a SDoH framework to research on CRC screening behaviors. The findings of this scoping review will provide a comprehensive overview of the SDoH associated with CRC screening behaviors among community residents in China. By mapping the range, frequency, and contextual meanings of these factors, this review will help identify which SDoH are most consistently linked to screening participation and follow-up adherence.

The findings will carry important implications for public health practice and community-level interventions. First, they will provide evidence to support the development of targeted strategies by health authorities, highlighting modifiable social barriers and facilitators within specific subpopulations. Second, the results will assist primary care providers and public health practitioners in identifying priority groups during the implementation of CRC screening programs, thereby enhancing the precision of resource allocation and improving coverage while reducing potential inequities in service delivery. Third, this review will contribute to the localization of SDoH measurement by synthesizing how these determinants have been defined and assessed in existing Chinese studies, offering a foundation for developing culturally adapted assessment tools.

**Systematic review registration:** Registered in OSF on April 23, 2025 (osf.io/v3mq7/)

## Introduction

Colorectal cancer (CRC) is one of the major malignant tumors threatening the health of the Chinese population. In 2022, China reported 517,106 new CRC cases and 240,010 deaths, accounting for approximately 26.9% and 26.5% of the global burden, respectively^[1]^.According to statistical data released by China’s National Cancer Center in 2024, CRC ranked second in cancer incidence and fourth in cancer-related mortality in China in 2022^[2]^,highlighting CRC as an urgent public health concern.A substantial body of evidence has confirmed that standardized CRC screening is an effective strategy to reduce both incidence and mortality rates, serving as a critical tool for early detection and early intervention of cancer^[3, 4]^.Four randomized trials assessing the effect of fecal occult blood testing (FOBT)-based screening found a 25% reduction in CRC mortality among individuals who participated in screening programs^[5]^.Long-term follow-up studies have shown that colonoscopy can reduce CRC incidence by approximately 40% and mortality by about 60%^[6, 7]^,thereby significantly decreasing the disease burden.Since 2012, China’s National Health Commission has launched a series of early detection and treatment initiatives for CRC, promoting community-based screening programs nationwide^[8]^. However, despite these efforts, screening coverage and follow-up adherence remain critically low in most regions.In 2020, the estimated coverage rate of organized CRC screening among individuals aged 40–74 was only 2.7%^[9]^.Among high-risk populations, the colonoscopy participation rate was merely 14%^[10]^,Even in Shanghai, one of China’s most developed regions, a large-scale community screening program revealed that only about 21.6% of the target population completed initial screening using the fecal immunochemical test (FIT) and high-risk factor questionnaire (HRFQ), and only 17.5% of those who tested positive underwent follow-up colonoscopy ^[11]^.Similarly, a CRC screening program in Chongqing reported that among 41,315 individuals who tested positive in preliminary screening, only 19.02% proceeded to the recommended colonoscopy^[12]^.Compared with the 60–70% CRC screening participation rates and 65–80% colonoscopy completion rates observed in high-income countries^[13]^,these data highlight significant implementation and uptake challenges in China. Multiple factors influence whether individuals participate in screening. These include not only biomedical factors such as age, sex, and family history^[14]^,but also non-medical factors such as educational attainment, income level, living environment, and access to medical services^[15-17]^.These are collectively categorized under the framework of SDoH, which refers to the conditions in which people are born, grow, work, live, and age, and their broader social and economic environments^[18]^.Multiple international studies have demonstrated that CRC screening behavior is profoundly influenced by various SDoH, including income level, accessibility of medical resources, educational background, residential environment, and social support networks^[15-17]^.Among these, low income and limited access to healthcare services are identified as the most prominent barriers. Populations experiencing economic hardship often face heightened health risks due to insufficient insurance coverage and limited access to health facilities ^[16, 19]^.Furthermore, disparities in education and inadequate social support—such as the absence of patient navigators or assistance from community health workers—can further reduce screening uptake and adherence among vulnerable groups^[20, 21]^.

To date, no systematic review or scoping synthesis has examined the impact of social determinants of health (SDoH) on colorectal cancer (CRC) screening behavior in China. Unlike the U.S. Department of Health and Human Services, which has established a standardized framework defining five core SDoH domains—economic stability, education access and quality, healthcare access and quality, neighborhood and built environment, and social and community context^[22]^—China lacks a comparable classification system. Existing studies are fragmented and lack comprehensive synthesis. To confirm this gap, we conducted a thorough bilingual literature search across Chinese and international databases, including CNKI, Wanfang Data, PubMed, Web of Science, Scopus, and Google Scholar. As of April 23, 2025, no published review or systematic synthesis specifically addressing the relationship between SDoH and CRC screening in China was identified.

Therefore, the main objective of the study is to systematically identify and synthesize existing evidence on SDoH associated with CRC screening participation among community residents in China, including their conceptual definitions and the instruments or measures used to assess these social determinants.

### Review Questions

This scoping review will address the following research questions:

1. What SDoH may be associated with CRC screening behavior among community residents in China?
2. What are the contextual meanings and interpretations of these SDoH within the local Chinese setting?
3. What types of questions or measurement tools have been used to investigate or assess these SDoH?

### Inclusion criteria

#### Participants

The target population of this scoping review comprises community-dwelling adults aged 40 years and older residing in mainland China, including both the general population (those eligible for initial screening) and individuals identified as high risk for CRC who require follow-up colonoscopy. High-risk individuals are defined as those who screen positive during the initial assessment using CRC risk stratification tools and/or FOBT.

#### Concept

According to the World Health Organization (WHO), SDoH refer to non-medical factors that influence health outcomes, including the conditions in which people are born, grow, live, work, and age, as well as broader social forces and institutional structures ^[18]^.In this study, we focus specifically on SDoH factors that are closely associated with CRC screening behavior among community residents in China.

#### Context

the community setting in China

#### Types of sources

Eligible studies include quantitative studies (e.g., cohort studies, cross-sectional studies, case-control studies), qualitative studies (e.g., interviews, focus groups, ethnographic studies), and mixed-methods studies combining quantitative and qualitative approaches.

## Methods

This study will be conducted in accordance with the JBI methodology for scoping reviews^[23]^.The review process will strictly adhere to this study protocol. Any deviations from the protocol will be clearly documented and justified in the Methods section.

### Inclusion Criteria

1. Studies focusing on community-dwelling residents in China aged 40 years and above.
2. Studies examining non-medical factors and their relationship with CRC screening behaviors.
3. Studies conducted in community settings in China, including urban and rural environments.
4. Eligible studies include quantitative studies (e.g., cohort studies, cross-sectional studies, case-control studies), qualitative studies (e.g., interviews, focus groups, ethnographic studies), and mixed-methods studies combining quantitative and qualitative approaches, published as Peer-reviewed journal articles, conference papers, and grey literature.
5. Studies published in English or Chinese.
6. Published after January 1, 2012, which marks the large-scale initiation of community-based CRC screening programs by China’s health authorities^[24]^.

### Exclusion Criteria

1. Studies focusing on non-community-dwelling populations (e.g., hospitalized patients, institutionalized individuals).
2. Studies where the majority of participants are under 40 years of age.
3. Studies conducted outside China.
4. Studies focusing solely on clinical or medical determinants (e.g., genetic factors) without reference to SDoH.
5. Studies conducted in non-community settings (e.g., hospitals, clinics) unless they explicitly address community-based CRC screening programs.
6. Reviews, editorials, opinion pieces, or studies lacking empirical data (e.g., no primary or secondary data collection).
7. Studies that do not report on SDoH and their relationship with CRC screening.
8. Studies published in languages other than English or Chinese.

### Search Strategy

The search strategy will combine both subject headings and free-text terms, with search terms structured around three key components: population, screening, and influencing factors. Separate search strategies will be developed for English and Chinese literature based on the JBI scoping review methodology^[25]^.

#### English-language search

An initial search will be conducted in PubMed and Embase using core concepts such as ” colorectal cancer,” ” screening,” ” social determinants,” and ” China.” The titles, abstracts, and keywords from the preliminary results will be analyzed using VOSviewer software (developed by the Centre for Science and Technology Studies, CWTS, at Leiden University) to identify frequently co-occurring terms and expand the list of relevant keywords. Based on the frequency and relevance of terms, an optimized search strategy will be developed (see Appendix 1). A subsequent full search will then be performed in PubMed, Embase, Web of Science, Scopus, and Google Scholar. Reference lists of included studies will be screened to identify additional potentially relevant studies.

#### Chinese-language search

An initial search will be conducted in two major Chinese databases—China National Knowledge Infrastructure (CNKI) and Wanfang Data. Core concepts will include terms such as ” colorectal cancer,” ” screening,” ” colonoscopy,” ” factors,” ” social determinants,” ” analysis,” ” community.” As with the English literature search, VOSviewer software will be used to analyze keyword frequencies from the retrieved literature, and the search strategy will be refined accordingly (see Appendix 1).

A second round of searches will then be conducted in CNKI and Wanfang using the optimized strategy. In addition, reference lists of included studies will be manually screened to identify further relevant publications.Given that most gray literature on this topic in mainland China is typically published in CNKI or Wanfang, we determined that further searches for gray literature would not be necessary.

The search for both Chinese and English literature will focus on publications from January 1, 2012, to April 23, 2025, aligning with the key period of policy implementation in China. If necessary, the researchers will contact the authors of relevant key studies to obtain additional information.

#### Study/Source of evidence selection

After completing the search, all records that meet the preliminary inclusion criteria will be imported into NoteExpress reference management software (Tongfang Knowledge Network, Beijing, China; Tongji University Library version). Duplicate records will be removed using a combination of automated and manual processes to generate a set of unique references for subsequent screening.

Two researchers (FX and YW), both familiar with the subject matter, will independently screen the titles and abstracts of the retrieved articles to exclude clearly irrelevant studies. Any article deemed potentially relevant by either reviewer will be retained for full-text screening.Full texts of the retained articles will then be retrieved and independently assessed by the same two researchers against the predefined inclusion criteria based on the PCC framework. Before the formal screening process begins, a pilot test will be conducted on the first 50 retrieved articles. This pilot screening of titles and abstracts will be used to clarify and harmonize the inclusion and exclusion criteria, ensuring consistency in their interpretation and application across reviewers.A weekly meeting will be held during the screening phase to discuss any discrepancies or ambiguous cases. Once an inter-reviewer agreement of over 90% is achieved in the pilot phase, the full screening process will proceed. In cases where disagreement persists and cannot be resolved through discussion, a third reviewer (DY) will act as an arbitrator. Final inclusion or exclusion decisions will be made through group discussion.

The study selection process will be presented in a PRISMA-ScR flow diagram, which will report the following stages:the total number of records retrieved;the number of records remaining after deduplication;the number of studies screened based on titles and abstracts;the number of full-text articles assessed for eligibility; and the final number of studies included in the review.A detailed log will be maintained to record all excluded full-text articles, along with explicit reasons for their exclusion (e.g., population mismatch, irrelevant concept, non-research article, etc.). These records will be securely stored in electronic format to ensure transparency and auditability of the review process. This information will be available upon request from the authors.

#### Data extraction

A standardized data extraction form will be developed to extract information from all studies included in the final review, systematically collecting and organizing relevant information. Two reviewers will independently extract key data from the included studies and cross-check each other’s results to ensure accuracy. Prior to full-scale extraction, a pilot extraction will be conducted on a small subset of studies to test and refine the data extraction form’s usability and comprehensiveness, with revisions made as necessary based on feedback from this phase.

The data items to be extracted include, but are not limited to:

First author’s name, year of publication, study location (province/city), community type (urban or rural), study design (e.g., cross-sectional, case-control, cohort study), specific characteristics of study participants (e.g., sample size, age range, gender distribution), screening methods used (e.g., community-based initial screening, follow-up colonoscopy uptake), SDoH related to screening behaviors, relationship between these SDoH and CRC screening behaviors, the contextual meanings of these factors in the Chinese setting (for qualitative studies only), and the specific survey questions or instruments used to measure them (for quantitative studies only).

After data extraction, the two reviewers will cross-check all entries. Any discrepancies will be resolved by reviewing the original articles to reach consensus; unresolved issues will be referred to a third team member for adjudication.

All verified data will be systematically compiled and securely stored in structured databases for subsequent analysis and audit purposes.

#### Data analysis and presentation

China has not yet established a nationally unified classification system or research paradigm for SDoH. Given this gap, the current review will adopt the Healthy People 2030 framework as an initial analytical structure.However, to ensure contextual relevance, we will also allow for flexible categorization of social factors that are repeatedly reported in the literature but not clearly aligned with the five established domains. These factors will be grouped under an additional category labeled ” Other”. For example, long-term migrant work status, if consistently reported as relevant to CRC screening behavior yet not directly attributable to the existing domains, will be classified under this supplementary category.

For each SDoH domain, we will present the identified influencing factors separately according to their relevance to the initial screening stage and the follow-up screening stage of CRC screening, or exert an impact on both stages. The initial screening stage refers to non-invasive preliminary screening procedures such as the FIT and HRFQ. The follow-up screening stage refers to further diagnostic procedures for individuals with positive results in the initial screening, such as colonoscopy.

The results of this review will be presented using a combination of narrative descriptions and visual formats such as tables and diagrams. A detailed narrative synthesis will be provided to explain how various categories of social determinants influence CRC screening behaviors. In addition, summary tables will be constructed to organize and present key findings (see Table Example 1). These tables will help readers to quickly identify and understand the characteristics of different social factors and their roles in CRC screening behavior.

**Table Example 1.**
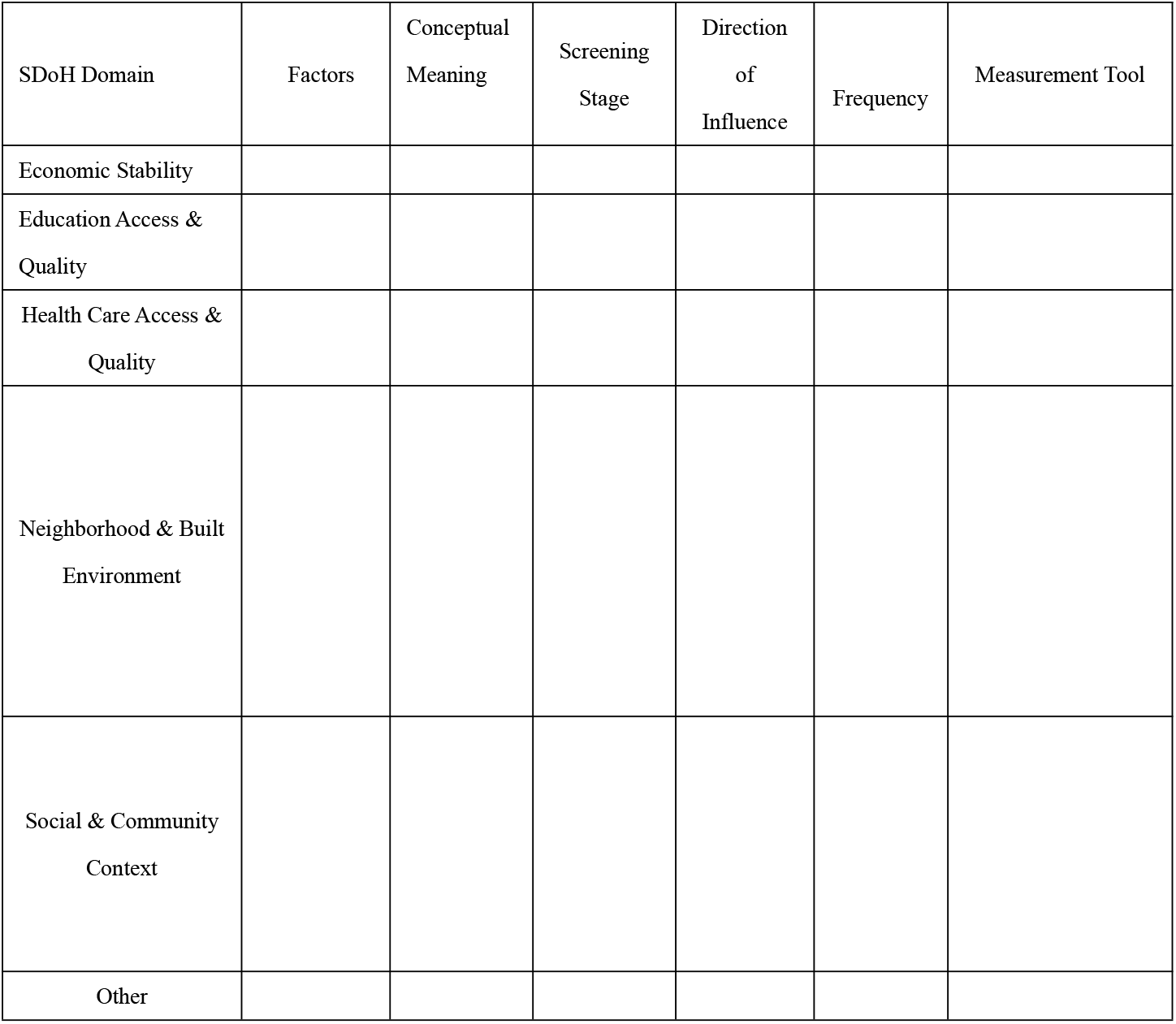
Summary of SDoH Factors Influencing CRC Screening.

## Data Availability

All data produced in the present work are contained in the manuscript

## Acknowledgements

Nil

## Funding

This research was funded by the Shanghai Municipal Health Commission Health Policy Research Project (Grant No. 2023HP28&2023HP71), Shanghai Leading Talents Program (Grant No. YDH-20170627), and Discipline Leader Advancement Program of Yangpu District Central Hospital (Ye2202103).

## Declarations

The authors declare no conflict of interest

## Author contributions

Conceptualization, F.X.; Methodology, F.X. ;L.W.and Y.W.; Data curation, F.X. and Y.W.; Formal analysis, F.X.; Funding acquisition, H.J. and D.Y.; Project administration, D.Y.; Resources, Y.W.;L.W.and H.J.; Supervision, D.Y.; Validation, F.X.; Writing—original draft, F.X.; Writing—review and editing, F.X., Y.W.,L.W.,H.J., D.Y.. All authors have read and agreed to the published version of the manuscript.

## Appendices

### Appendix I: Search strategy

#### Quantitative/Qualitative Studies (English)

PubMed

(” Colorectal Neoplasms” [Mesh] OR ” colorectal cancer” [tiab] OR ” colon cancer” [tiab] OR ” rectal cancer” [tiab] OR ” bowel cancer” [tiab] OR CRC[tiab])

AND

(” Mass Screening” [Mesh] OR ” Early Detection of Cancer” [Mesh] OR screening[tiab] OR ” early detection” [tiab] OR ” screening uptake” [tiab] OR ” screening participation” [tiab] OR ” screening adherence” [tiab] OR ” screening compliance” [tiab] OR ” screening rate” [tiab] OR ” screening behavior” [tiab])

AND

(” Social Determinants of Health” [Mesh] OR ” Socioeconomic Factors” [Mesh] OR ” Healthcare Disparities” [Mesh] OR ” Health Services Accessibility” [Mesh] OR ” Educational Status” [Mesh] OR ” social determinant*” [tiab] OR ” socioeconomic” [tiab] OR education[tiab] OR income[tiab] OR poverty[tiab] OR ” healthcare access” [tiab] OR ” social support” [tiab] OR ” community context” [tiab] OR urban[tiab] OR rural[tiab] OR ” health equity” [tiab])

AND

(” China” [Mesh] OR China[tiab] OR Chinese[tiab]) NOT

(” Hong Kong” [tiab] OR Taiwan[tiab] OR ” Chinese American” [tiab]) AND

(English[lang])

Embase

(‘colorectal cancer’/exp OR ‘colon cancer’:ti,ab OR ‘rectal cancer’:ti,ab OR ‘colorectal neoplasm*’:ti,ab OR ‘bowel cancer’:ti,ab OR CRC:ti,ab)

AND

(‘screening’/exp OR ‘early detection’:ti,ab OR ‘screening uptake’:ti,ab OR ‘screening participation’:ti,ab OR ‘screening adherence’:ti,ab OR ‘screening behavior’:ti,ab)

AND

(‘social determinant*’:ti,ab OR ‘socioeconomic’:ti,ab OR ‘education’:ti,ab OR ‘income’:ti,ab OR ‘poverty’:ti,ab OR ‘healthcare access’:ti,ab OR ‘social support’:ti,ab OR ‘community context’:ti,ab OR ‘urban’:ti,ab OR ‘rural’:ti,ab OR ‘health inequality’:ti,ab)

AND

(‘China’/exp OR China:ti,ab OR Chinese:ti,ab) NOT

(‘Hong Kong’:ti,ab OR Taiwan:ti,ab OR ‘Chinese American*’:ti,ab)

Web of Science

TS=(” colorectal cancer” OR ” colon cancer” OR ” rectal cancer” OR ” colorectal neoplasm*” OR ” bowel cancer” OR CRC)

AND

TS=(” screening” OR ” early detection” OR ” screening participation” OR ” screening adherence” OR ” screening behavior”)

AND

TS=(” social determinant*” OR socioeconomic OR education OR income OR poverty OR ” healthcare access” OR ” social support” OR ” community context” OR urban OR rural OR ” health disparity” OR ” health equity”)

AND

TS=(China OR Chinese)

NOT

TS=(” Hong Kong” OR Taiwan OR ” Chinese American*”) AND

LA=(English)

Scopus

TITLE-ABS-KEY(” colorectal cancer” OR ” colon cancer” OR ” rectal cancer” OR ” colorectal neoplasm*” OR ” bowel cancer” OR CRC)

AND

TITLE-ABS-KEY(” screening” OR ” early detection” OR ” screening uptake” OR ” screening participation” OR ” screening adherence” OR ” screening behavior”)

AND

TITLE-ABS-KEY(” social determinant*” OR socioeconomic OR education OR income OR poverty OR ” healthcare access” OR ” social support” OR ” community context” OR urban OR rural OR ” health equity”)

AND

TITLE-ABS-KEY(” China” OR ” Chinese”)

AND NOT

TITLE-ABS-KEY(” Hong Kong” OR ” Taiwan” OR ” Chinese American*”) AND

(LIMIT-TO (LANGUAGE, ” English”))

Google Scholar

(” colorectal cancer screening” AND China AND ” community” AND (” social determinants” OR socioeconomic OR education OR income OR poverty OR urban OR rural))

-” Hong Kong” -Taiwan -” Chinese American”

#### Quantitative/Qualitative Studies (Chinese)

##### China National Knowledge Infrastructure (CNKI)

TKA=(” 结直肠癌” OR ” 大肠癌” OR ” 结肠癌” OR ” 直肠癌” OR ” 肠癌” OR ” 肠道肿瘤”) (” colorectal cancer” OR ” colon cancer” OR ” rectal cancer” OR ” large bowel cancer” OR ” intestinal cancer” OR ” intestinal tumor”)

AND

TKA=(” 筛查” OR ” 早期发现” OR ” 早诊早治” OR ” 初筛” OR ” 复筛” OR ” 粪便潜血” OR ” FOBT” OR ” FIT” OR ” HRFQ” OR ” 高危因素问卷” OR ” 参与率” OR ” 依从性” OR ” 顺应

性” OR ” 肠镜”)(” screening” OR ” early detection” OR ” early diagnosis and treatment” OR ” initial screening” OR ” rescreening” OR ” fecal occult blood” OR ” FOBT” OR ” FIT” OR ” HRFQ” OR ” high-risk factor questionnaire” OR ” participation rate” OR ” compliance” OR ” adherence” OR ” colonoscopy”)

AND

TKA=(” 因素” OR ” 分析” OR ” 社会支持” OR ” 社会环境” OR ” 经济” OR ” 收入” OR ” 教育” OR ” 医保” OR ” 文化” OR ” 城乡差距” OR ” 社会地位”)(” factors” OR ” analysis” OR ” social support” OR ” social environment” OR ” economy” OR ” income” OR ” education” OR ” medical insurance” OR ” culture” OR ” urban-rural disparity” OR ” social status”)

AND

TKA=(” 社区” OR ” 居民” OR ” 基层”)(” community” OR ” residents” OR ” primary level”)

Wanfang Data

Subject=(” 结直肠癌” OR ” 大肠癌” OR ” 结肠癌” OR ” 直肠癌” OR ” 肠癌” OR ” 肠道肿瘤”)

(” colorectal cancer” OR ” colon cancer” OR ” rectal cancer” OR ” bowel cancer” OR ” colonic cancer” OR ” intestinal tumor”)

AND

Subject=(” 筛查” OR ” 早期发现” OR ” 早诊早治” OR ” 初筛” OR ” 复筛” OR ” 粪便潜血” OR ” FOBT” OR ” FIT” OR ” HRFQ” OR ” 高危因素问卷” OR ” 参与率” OR ” 依从性” OR ” 顺应性”

OR ” 肠镜”)(” screening” OR ” early detection” OR ” early diagnosis and treatment” OR ” initial screening” OR ” rescreening” OR ” fecal occult blood” OR ” FOBT” OR ” FIT” OR ” HRFQ” OR ” high-risk factor questionnaire” OR ” participation rate” OR ” adherence” OR ” compliance” OR ” colonoscopy”)

AND

Subject=(” 因素” OR ” 分析” OR ” 社会支持” OR ” 社会环境” OR ” 经济” OR ” 收入” OR ” 教育” OR ” 医保” OR ” 文化” OR ” 城乡差距” OR ” 社会地位”)(” factors” OR ” analysis” OR ” social support” OR ” social environment” OR ” economy” OR ” income” OR ” education” OR ” medical insurance” OR ” culture” OR ” urban-rural gap” OR ” social status”)

AND

Subject=(” 社区” OR ” 居民” OR ” 基层”)(” community” OR ” residents” OR ” primary level”)

The search in all databases will be limited to publications from **January 1, 2012, to April 23,** Search conducted on: 23,May, 2025

## Notes

### Competing Interest Statement

The authors have declared no competing interest.

